# Translation and Validation of the Brazilian – Portuguese version of Quality of Life on Vascular Access Devices Specific Questionnaire in Chemotherapy patients

**DOI:** 10.1101/2025.01.29.25320594

**Authors:** Bruno Jeronimo Ponte, Carolina Carvalho Jansen Sorbello, Ricardo Ferreira Mendes de Oliveira, Maria Fernanda Cassino Portugal, Andressa Cristina Sposato Louzada, Marcelo Fiorelli Alexandrino da Silva, Lucas Lembrança Pinheiro, Cynthia de Almeida Mendes, Nelson Wolosker

## Abstract

1.

**Background:** The use of long-term devices for chemotherapy in neoplastic diseases is very common. The CAVA trial was an extensive study that prospectively evaluated more than a thousand patients undergoing chemotherapy and randomized them to three groups: patients using PICC, Port or Hickmann-type catheters. For this study, a specific questionnaire was elaborated, called the *Venous access device quality of life questionnaire* (QolVAD), the aim of which is to assess the impact of the device (Hickmann type, PICC or thoracic Port) on the patients’ routine. However, there is no Brazilian – Portuguese version available, blunting its use in Brazilian patients.

**Objective:** The objective of this study is to translate and validate de CAVA Trial QolVAD to the Brazilian – Portuguese, making possible its use in Brazilian population.

**Methods:** One hundred sixty-seven patients with long term vascular access devices for chemotherapy participated in the study. After translation and retranslation, construct validity was analyzed by identifying correlation between QolVAD and European Quality of Life Questionnaire (EuroQol). To determine the reliability, internal consistency and test-retest with at least 7 days interval between 2 questionnaire applications were calculated.

**Results:** The results showed a good internal and external consistency of the QolVAD. When comparing to EuroQol, significant correlations were found after comparing both questionnaires (r −0,658 and p < 0,001).

**Conclusion:** The Brazilian – Portuguese version of the Quality-of-Life Vascular Access Device has presented good clinimetric properties and has shown to be applicable to the Brazilian population.

## 2. Introduction

The use of long-term vascular devices for chemotherapy in cancer treatments is widespread. ^1^ The National Cancer Institute estimated that there would be approximately 704.000 new cancer cases in Brazil in 2023. ^2^

When chemotherapy needs to be administered intravenously, it can be done through either peripheral or dedicated vascular access. ^3^ The second category includes peripherally inserted central catheters (PICCs), partially implantable tunneled long-term central catheters (Hickmann), and fully implantable tunneled central catheters (PORTs). ^4^ These devices are particularly beneficial for long-term intravenous therapy because they deliver medication directly into central veins. This reduces the risk of complications, such as phlebitis and extravasation, that can occur in smaller veins when using vesicant chemotherapy drugs. ^5^

While numerous studies have examined complication outcomes of catheter types, there remains a lack of literature regarding the quality of life related to each catheter modality. ^6–9^

The CAVA trial was a comprehensive study that prospectively evaluated over a thousand patients undergoing chemotherapy.^10^ These patients were randomized into three groups based on the type of venous access device they used: PICC, Port, or Hickmann-type catheters. ^5,10^ The authors found that patients using PORT-type devices experienced fewer catheter-related complications. ^5^ To assess the impact of these devices on patients’ daily lives, the study utilized a specific questionnaire called the Venous Access Device Quality of Life Questionnaire (QolVAD). An evaluation of the psychosocial aspects related to the catheters used by CAVA trial patients revealed a preference for PORT-type devices. However, this preference was not universal among all participants. ^11^

In another study focused exclusively on patients using PICC devices, the authors determined that most patients considered using the device to be a positive experience that does not affect their quality of life. ^12^ Additionally, a second study focused specifically on patients with PICCs, while categorizing them based on the type of primary tumor, concluded that the device has minimal effect on a patient’s quality of life. Most issues reported were related to the underlying disease rather than the catheter itself. ^13^

In a prospective cohort of 35 patients receiving brachial insertion for PICC-type catheters, 94.3% of respondents would recommend the device to others. Most patients reported no harm to their daily routines or feelings of anxiety about the device. ^14^

High-quality data from randomized studies with high statistical power are essential for guiding clinical practices and decisions regarding vascular access in long-term intravenous therapies. However, we currently lack a translated and validated tool in our language to assess the quality of life for patients with long-term catheters.

The goal of this study was to translate and validate the QolVAD used by the CAVA trial research group ^5,10^ into Brazilian Portuguese.

## 3. Methods

This study aimed to translate and validate the CAVA Trial questionnaire into Brazilian Portuguese. The research was conducted with patients undergoing chemotherapy through long-term venous access during their cancer follow-up at a tertiary complexity oncological center.

This study followed the principles outlined in the Helsinki Declaration and received approval from the institution’s Ethics Committee under protocol number 59982622.3.0000.0071. Before participating, all patients received comprehensive information about the study’s objectives and procedures, and informed consent was obtained from each participant.

All data was collected was handled confidentially and stripped of any identifying information (de-identified data).

### Translation

The standard “forward-backward” method was used to translate the questionnaire developed by the authors of the CAVA Trial from English to Portuguese.

Two health professionals fluent in English and whose native language was Brazilian Portuguese were selected for the task. They translated the items and answers or the questionnaire to create a provisional version. This initial version was then tested on two voluntary patients, and adjustments were made to start the re-translation process.

Next, two independent professionals with the same qualifications re-translated the provisional version back into English. This step was taken to assess its compatibility with the original English version. The re-translated questionnaire was then culturally adapted and renamed the Revised Provisional Version.

Following this, two members of the research group, both vascular surgeons, checked the Revised Provisional Version to ensure it effectively assessed the necessary elements for analyzing patients’ quality of life when using catheters for chemotherapy.

The researchers compared the various versions with the original text, identifying and correcting any discrepancies. A consensus version was then formulated, with careful attention paid to preserving the semantic equivalence and ensuring the vocabulary was simple and direct.

### Sample

A convenience sample of 180 patients of both genders in this study, which was conducted in an oncological outpatient setting. The inclusion criteria were patients over the age of 18 undergoing chemotherapy via a PICC or PORT implanted in the last 30 days and who agreed to take part in this study by signing an informed consent form. Exclusions were made for patients under 18 years old, those who had been using venous devices for less than 30 days prior to the administration of the questionnaire, patients who died between questionnaire reapplications, and individuals who underwent catheter changes within that time frame.

The included participants were asked about aspects related to the quality of life-related to the vascular devices they were currently using, whether PICC or PORT. The data was collected using two assessment tools: a questionnaire created by the authors of the CAVA Trial called Quality of Life - Vascular Access Device (QolVAD), and the European Quality of Life Questionnaire (EuroQol, EQ-5D-5L).

All subjects received comprehensive information about the objectives and procedures of the study. Those who voluntarily agreed to participate in the study signed an informed consent form.

### Validation

To analyze the validity of the QoLVAD construct, we compared data obtained from its 16 questions with the EQ-5D-5L index value. The data from the QolVAD questionnaire were collected through interviews conducted by a single examiner. To determine the final QolVAD score, the average of the responses was calculated and applied a multiplier of 10, resulting in scores ranging from 10 to 40. Given the scoring system, higher QolVAD scores indicate a worse quality of life for the patients.

The EuroQol has 5-domain version with 5 response options per question (EQ-5D-5L), which has been translated into Brazilian Portuguese.^15^ Each country has a predefined value set used to calculate the final index score, making it possible to highlight the differences between each nation. Since a value set for the EQ-5D-5L is not yet available in Brazil, we calculated the index score using the value from the United States, as recommended by the EuroQol organization. ^16^ In contrast to QolVAD, the EuroQol index score ranges from a maximum of 1 (indicating the best quality of life) to lower numbers, which represents worse quality of life.

The QolVAD questionnaire was applied to patients with all 16 questions, and their answers ranged from 1 (does not affect quality of life) to 4 (affects it a lot). A non-apply category (0) was included, as the number of patients did not correlate with certain questions (e.g., regarding difficulties in driving a car – applicable only when the patient drives).

### Internal Consistency

To analyze the reliability of the QolVAD, a sub-sample of 46 patients was interviewed on two different occasions: occasion 1 and occasion 2, at least 7 days apart and no more than 30 days. At both times, the interviews were conducted by the same evaluator.

The inter-observer analysis was carried out by interviewing a total of 31 patients, with the Qol VAD being applied at two different times by two different observers within a maximum time interval of 7 days.

### Statistical Analysis

The demographic analysis was conducted by calculating the means and medians of the assessed parameters: age, sex, and body mass index (BMI).

The association between QolVAD and EQ-5D-5L was analyzed using Spearman’s rank correlation coefficient. A coefficient greater than 0.5 was considered sufficient to validate the questionnaire, with a statistical significance set at *p <0*,*05*.

We compared the qualitative variants from the questionnaire using Kendall’s Tau correlation coefficient for internal validation and inter-observer evaluation analysis. A coefficient below 0,5 was considered weak and above 0,5 a moderate correlation, with statistical significance also set at *p<0*,*05*.^17^

## 4. Results

Among the 180 patients established and invited, only 167 agreed to take part in the study. Table 1 presents the demographic characteristics of the interviewed patients. The distribution of men and women was approximately equal. Most patients were in their sixties and were not considered obese.

**Table 1:**
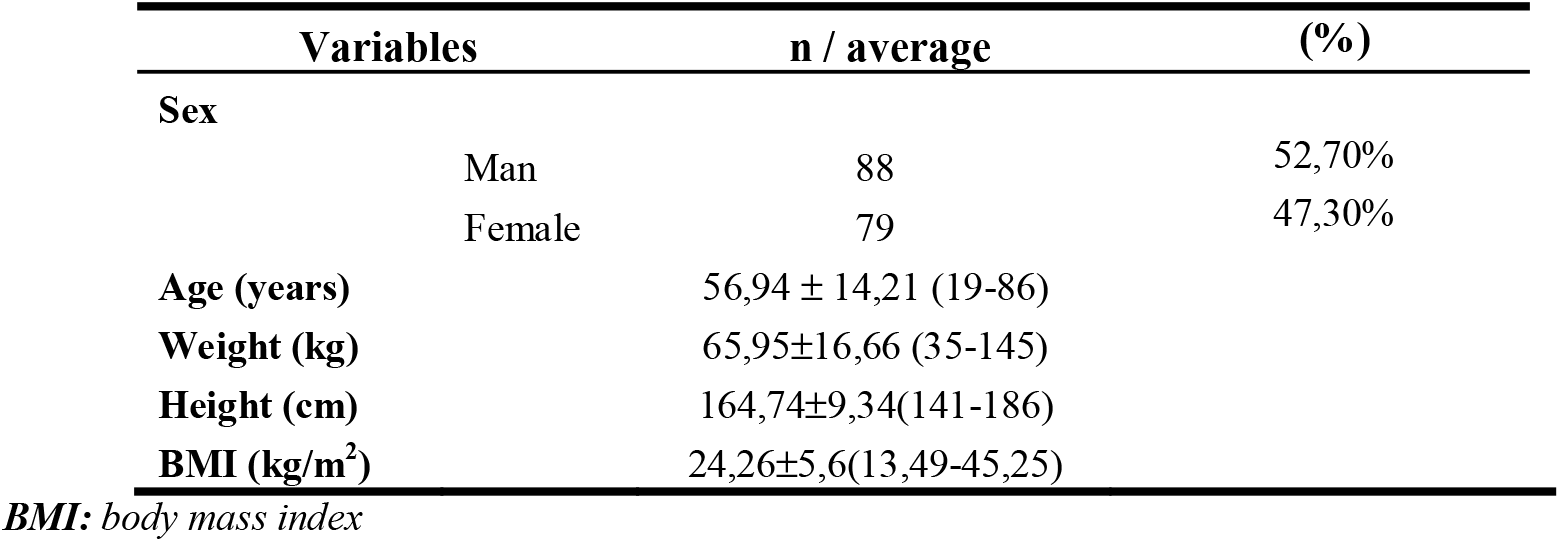
Characteristics of the patients.

Figure 1 illustrates the Spearman correlation coefficients between the QolVAD and EQ-5D-5L questionnaires. The negative coefficient occurred due the different parameters used by the two questionnaires. In QolVAD, higher scores indicate a poorer quality of life, whereas higher scores in EuroQol reflects a better quality of life. The correlation was found to be negative, and statistically significant, with a coefficient of r = −0.658 (*p<0*.*001*).

**Figure 1:**
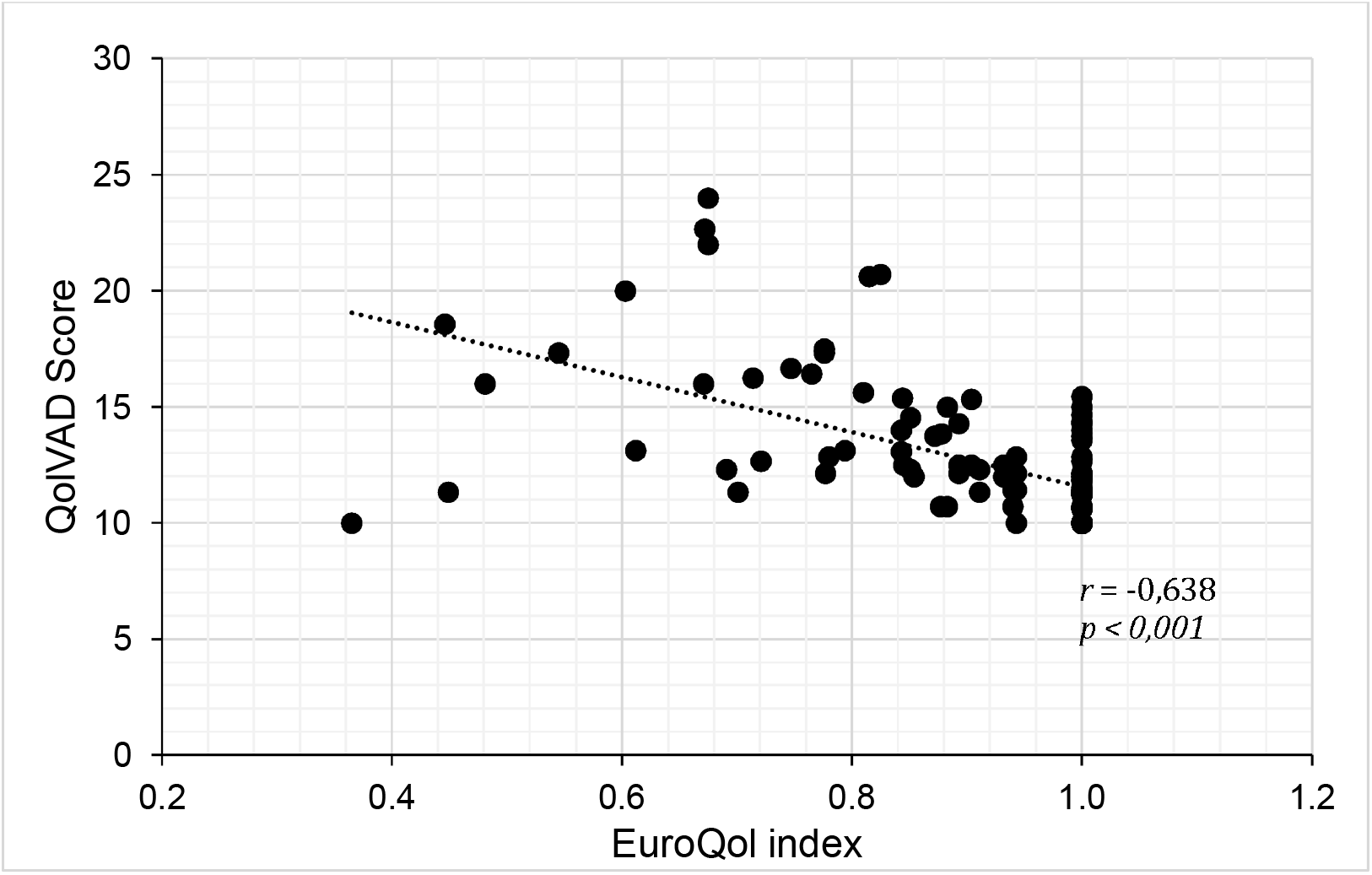
Scatter plot demonstrating the correlation between QolVAD scores and EuroQol index values per questionnaire applied using Spearman’s coefficients correlation.

Table 2 presents data on the reliability of the QolVAD questionnaire. The results reflect responses by the same interviewer on two separate occasions, which were analyzed using Kendall’s Tau B correlation test. Questions 1, 2, 3, and 8 exhibited no variability in responses, resulting in an absolute agreement between the two moments the QolVAD questionnaire was applied. Although a total of 46 patients completed the questionnaires, only 26 patients responded to question 9, which limited the significance of the analysis. All other questions demonstrated a moderate correlation and were statistically significant.

**Table 2:** Internal Validation. Kendall’s Tau rank correlation coefficient was used to determine the internal validation. *: Absolute agreement between the questionnaires applied by the same observer.

| Qol VAD Question | R | P |
| --- | --- | --- |
| 1 | 1 | * |
| 2 | 1 | * |
| 3 | 1 | * |
| 4 | 0,609 | <0,001 |
| 5 | 0,699 | <0,001 |
| 6 | 0,989 | <0,001 |
| 7 | 1 | <0,001 |
| 8 | 1 | * |
| 9 | 0,348 | 0,082 |
| 10 | 0,564 | <0,001 |
| 11 | 0,656 | <0,001 |
| 12 | 1 | <0,001 |
| 13 | 1 | <0,001 |
| 14 | 0,5 | <0,001 |
| 15 | 0,627 | <0,001 |
| 16 | 0,693 | <0,001 |

For the inter-observer analysis, 31 out of 167 patients were interviewed by two observers at different times. Statistical analysis was carried out using Kendall’s Tau correlation coefficient due to the small sample size and minimal variability in the responses. The results are presented in Table 3. All questions demonstrated positive correlations. Specifically, Questions 1, 2, 3, 5, 8, 9, 10, 11, and 13 had the same answers with both observers, resulting in an analysis with absolute agreement. Although Question 16 was not statistically significant, it still exhibited a positive correlation with the sample.

**Table 3:** Inter-Observer validation. Observer 1 and 2 questionnaires were compared using Kendall’s Tau rank correlation coefficient. *: Absolute agreement between the questionnaires applied by the different observers.

| **QoIVAD Question** | **R** | **P** |
| --- | --- | --- |
| **1** | 1 | * |
| **2** | 1 | * |
| **3** | 1 | * |
| **4** | 0,462 | 0,016 |
| **5** | 1 | * |
| **6** | 0,822 | <0,001 |
| **7** | 0,631 | <0,001 |
| **8** | 1 | * |
| **9** | 1 | * |
| **10** | 1 | * |
| **11** | 1 | * |
| **12** | 1 | <0,001 |
| **13** | 1 | * |
| **14** | 0,522 | 0,001 |
| **15** | 0,694 | <0,001 |
| **16** | 0,334 | 0,062 |

## 5. Discussion

This study aimed to translate and validate the questionnaire produced by the CAVA Trial (QolVAD) authors in a Brazilian sample. The objective was to create a specific questionnaire for assessing the quality of life in patients using long-term catheters for chemotherapy.

The QolVAD Questionnaire was developed by the authors of the CAVA Trial to specifically assess the change in the quality of life caused by PICC, PORT, and Hickmann catheters in patients undergoing chemotherapy. ^5,10^

The Portuguese version of the QolVAD was translated using the forward-backward methodology. ^18,19^ Few changes were made to the questionnaire to ensure cultural appropriateness, as the items proved to apply to the Brazilian context. When assessing patients’ work performance, we included individuals who reported engaging in domestic activities, as many patients were not working due to absence from work due to the cancer disease. Additionally, in the last question of the QolVAD, we also considered the impact of PICC catheters on the frequency of hospital visits for changing PICC-related dressings.

Several widely recognized questionnaires assess general quality of life, with the most frequently used being the SF-36 and EuroQol - both of which have already been translated and validated into Portuguese. ^15^ To carry out the analysis for external validation, the QolVAD and EuroQol questionnaires were compared. This questionnaire was chosen because it has a well-documented Portuguese version and is concise and easy to understand for the patients we approached. ^15^

The analysis between the QolVAD and EQ-5D-5L was done by comparing the average value of the QolVAD answers with the value of the EuroQol index. As Brazil does not currently have the set of values needed to calculate the EQ-5D-5L index, we used the set of values established by the United States, as recommended by EuroQol. ^16,20^ We found a negative coefficient, corroborating the construct analysis between the two questionnaires.

To validate inter-observer reliability, patients were interviewed by two different observers who had previously been trained for the assessment process. The questionnaires were compared using Kendall’s Tau - B test, which demonstrated that the questions on the QolVAD were positively consistent between the different observers. Question 16 was the only one that was not statistically significant, with a poor but positive correlation. It evaluates aspects of the quality of life of patients using catheters. We believe that the different times the surveys were administered may have influenced the results on this subjective question.

The internal validation analysis demonstrated a positive correlation between the responses the same observer obtained at different times. All the questions demonstrated a positive correlation. Question 9 addresses how the use of vascular access affects the job of these patients. As many of the institution’s cancer patients were away from work due to cancer treatment, many participants skipped this question. We also noticed that some patients stopped working because of their treatment between our interviews, and had different answers at different times when the questionnaire was applied.

Some limitations were encountered in validating the QolVAD. While collecting the questionnaires, it was noted that many patients did not answer specific QolVAD questions. In particular, questions about driving, using public transport, shopping, working, or engaging in physical activities were often left unanswered, as some patients did not participate in these activities. Consequently, these questions were categorized as “Does not Apply,” a designation not included in the original questionnaire, leading to reduced sample size for statistical comparison (as Question 9 in internal validation). Additionally, some patients experienced changes in their functional status and activities of daily living due to cancer and/or its treatment, which further impacted the statistical analysis by resulting in varying scores on the questionnaires at different times (as Question 16 on inter-observer reliability). Added to that, external validation was carried out by comparing an established score to the QolVAD questionnaire that is not described by the authors of the CAVA Trial, which weakens external validation statistically.

From our results, we believe that the translation and validation process of the QolVAD questionnaire into Brazilian Portuguese was satisfactory. It was found to have statistically significant correlation with other questionnaire (construct validation) and good reproducibility indices. Hence, we can safely say that the Brazilian Portuguese version of the QolVAD can be used in clinical practice to assess the impact of different vascular accesses for chemotherapy on patients’ quality of life.

## 6. Conclusion

The Brazilian Portuguese version of the QolVAD presents adequate validity and reliability indicators, which supports its application to Brazilian patients with long-term vascular access for chemotherapy.

## Data Availability

All data produced in the present study are available upon reasonable request to the authors

## References

1. Wolosker N, Yazbek G, Nishinari K, Malavolta LC, Munia MA, Langer M, et al. Totally implantable venous catheters for chemotherapy: experience in 500 patients. J Vasc Bras. 2017;(16):128–39.

2. Ministério da Saúde Instituto Nacional de Câncer José Alencar Gomes da Silva Ministério da Saúde Instituto Nacional de Câncer. 2023.

3. Zerati AE, Wolosker N, de Luccia N, Puech-Leão P. Totally implantable venous catheters: history, implantation technique and complications. Vol. 16, Jornal Vascular Brasileiro. Sociedade Brasileira de Angiologia e Cirurgia Vascular; 2017. p. 128–39.

4. Wolosker N, Yazbek G, Munia MA, Zerati AE, Langer M, Nishinari K. Totally implantable femoral vein catheters in cancer patients. European Journal of Surgical Oncology. 2004 Sep;30(7):771–5.

5. Wu O, McCartney E, Heggie R, Germeni E, Paul J, Soulis E, et al. Venous access devices for the delivery of long-term chemotherapy: The cava three-arm rct. Health Technol Assess (Rockv). 2021 Jul 1;25(47):7–102.

6. Patel GS, Jain K, Kumar R, Strickland AH, Pellegrini L, Slavotinek J, et al. Comparison of peripherally inserted central venous catheters (PICC) versus subcutaneously implanted port-chamber catheters by complication and cost for patients receiving chemotherapy for non-haematological malignancies. Supportive Care in Cancer. 2014;22(1):121–8.

7. Taxbro K, Hammarskjöld F, Thelin B, Lewin F, Hagman H, Hanberger H, et al. Clinical impact of peripherally inserted central catheters vs implanted port catheters in patients with cancer: an open-label, randomised, two-centre trial. Br J Anaesth. 2019 Jun 1;122(6):734–41.

8. Leiderman DBD, Souza KP, Binatti CET, Mendes C de A, Teivelis MP, Brito CF, et al. Arm mobilization provokes deformity of long-term indwelling ports implanted via the jugular vein. J Vasc Surg Venous Lymphat Disord. 2021 Jul 1;9(4):998–1006.

9. Pu Y Lou, Li ZS, Zhi XX, Shi YA, Meng AF, Cheng F, et al. Complications and Costs of Peripherally Inserted Central Venous Catheters Compared with Implantable Port Catheters for Cancer Patients: A Meta-analysis. Cancer Nurs. 2020 Nov 1;43(6):455– 67.

10. Moss JG, Wu O, Bodenham AR, Agarwal R, Menne TF, Jones BL, et al. Central venous access devices for the delivery of systemic anticancer therapy (CAVA): a randomised controlled trial. The Lancet. 2021 Jul 31;398(10298):403–15.

11. Ryan C, Hesselgreaves H, Wu O, Moss J, Paul J, Dixon-Hughes J, et al. Patient acceptability of three different central venous access devices for the delivery of systemic anticancer therapy: A qualitative study. BMJ Open. 2019 Jul 1;9(7).

12. Parás-Bravo P, Paz-Zulueta M, Santibañez M, Fernández-de-las-Peñas C, Herrero-Montes M, Caso-Álvarez V, et al. Living with a peripherally inserted central catheter: the perspective of cancer outpatients—a qualitative study. Supportive Care in Cancer. 2018 Feb 1;26(2):441–9.

13. Kang J, Chen W, Sun W, Ge R, Li H, Ma E, et al. Health-related quality of life of cancer patients with peripherally inserted central catheter: A pilot study. Journal of Vascular Access. 2017;18(5):396–441.

14. Fonseca IYI, Krutman M, Nishinari K, Yazbek G, Teivelis MP, Bomfim GAZ, et al. Brachial insertion of fully implantable venous catheters for chemotherapy: complications and quality of life assessment in 35 patients. Einstein (Sao Paulo). 2016 Oct 1;14(4):473–9.

15. Ferreira PL, Ferreira LN, Pereira LN. contribution for the Validation of the Portuguese Version of EQ-5D contributos para a Validação da Versão Portuguesa do EQ-5D [Internet]. Available from: https://www.actamedicaportuguesa.com

16. Van Reenen M, Janssen B, Stolk E, Boye KS, Herdman M, Kennedy-Martin M, et al. EQ-5D-5L User Guide: Basic information on how to use the EQ-5D-5L instrument [Internet]. Available from: https://euroqol.org/publications/user-guides.

17. Koo TK, Li MY. A Guideline of Selecting and Reporting Intraclass Correlation Coefficients for Reliability Research. J Chiropr Med. 2016 Jun 1;15(2):155–63.

18. Mendes Ritti-Dias R, Alberto Gobbo L, Grizzo Cucato G, Wolosker N, Jacob Filho W, Maria Santarém J, et al. Translation and Validation of the Walking Impairment Questionnaire in Brazilian Subjects with Intermittent Claudication.

19. Varella AYM, Fukuda JM, Teivelis MP, De Campos JRM, Kauffman P, Cucato GG, et al. Translation and validation of Hyperhidrosis Disease Severity Scale. Rev Assoc Med Bras. 2016 Dec 1;62(9):843–7.

20. Pickard AS, Law EH, Jiang R, Pullenayegum E, Shaw JW, Xie F, et al. United States Valuation of EQ-5D-5L Health States Using an International Protocol. Value in Health. 2019 Aug 1;22(8):931–41.

